# Circulating and Adipose Tissue Profiles of Fatty Acid Esters of Hydroxy-Fatty Acids in Women: Impact of Adiposity, Age, and Acute Exercise

**DOI:** 10.64898/2026.05.13.26352871

**Authors:** Lenka Rossmeislová, Viktor Šebo, Jan Gojda, Michal Koc, Marek Wilhelm, Martin Riečan, Tomáš Čajka, Jana Potočková, Jana Neubert, Eva Krauzová, Alejandro Ezequiel Harnichar, Ondřej Kuda, Michaela Šiklová, Martin Rossmeisl

## Abstract

**Objective:** Fatty Acid esters of Hydroxy-Fatty Acids (FAHFAs) are anti-diabetic and anti-inflammatory lipokines produced mainly by adipose tissue (AT). As exercise training enhances FAHFA levels, we investigated the impact of acute exercise (AE) and exercise-mimicking conditions on circulating and adipocyte FAHFA levels.

**Methods:** Clinical trial (NCT05572905) in 60 women, grouped by BMI (lean vs. obese) and age (young vs. older), was combined with *in vitro* experiments on human adipocytes. Following baseline characterization (body composition, VO_2max_, insulin sensitivity, AT/plasma FAHFAs), women underwent a cross-over AE and control interventions with repeated blood sampling for FAHFA analysis.

**Results:** In AT, lean and older women exhibited higher FAHFA levels than obese and young women, respectively; older women also showed a shift toward higher levels of 13/12-carbon-branched FAHFAs. Circulating FAHFA levels were similar across all groups and were not positively associated with insulin sensitivity, VO_2max_ or FAHFA levels in AT. Although AE increased circulating free fatty acids (FFA), plasma FAHFAs dropped in response to both AE and control interventions. In adipocytes, FAHFAs were unaffected by glucocorticoids but increased in response to lipolysis together with gene expression related to FFA oxidation (FAO). Nevertheless, blocking mitochondrial FAO partially mimicked the lipolytic effect, while peroxisomal inhibition synergistically boosted FAHFA lipolysis-driven production despite having no effect alone.

**Conclusions:** While adiposity and aging modulate FAHFA levels in AT, circulating levels remain stable and unaffected by AE, challenging subcutaneous AT as their primary systemic source. *In vitro*, FAHFA synthesis is driven by high FFA availability but limited by competing peroxisomal FAO.

## 1. Introduction

Adipose tissue (AT) is a rich source of signaling molecules modulating metabolic processes. Among these, fatty acid esters of hydroxy-fatty acids (FAHFAs), identified in 2014, represent a class of lipokines with potent insulin-sensitizing and anti-inflammatory properties [1, 2]. FAHFAs, especially palmitic acid esters of hydroxy-stearic acid (PAHSAs), have been shown to enhance glucose uptake in adipocytes, reduce hepatic glucose production, stimulate insulin secretion, and mitigate inflammation [3–6]. Nevertheless, the structural complexity of FAHFAs—arising from numerous ester combinations, branching, and chirality—means that most regioisomers beyond PAHSAs remain poorly understood [3, 7, 8].

Consistent with their beneficial effect on glucose homeostasis, FAHFA levels are characteristically decreased in obesity and insulin-resistant states [1, 9, 10]. Restoring FAHFA levels—whether through bariatric surgery or exercise training—has been associated with improved metabolic health [9, 11]. Importantly, the exercise-driven increase of FAHFAs in both AT and circulation [11] suggests an active role of subcutaneous AT (SAT) in exercise-induced adaptations.

Despite these findings, the impact of acute exercise on FAHFAs remains less understood. Nelson et al. reported a decrease in circulating FAHFAs following exercise in young trained individuals, who were lean, but not in subjects with overweight [12]. But it remains unknown how age affects this FAHFA dynamics, as it can influence AT function independently of adiposity. In addition, previous findings suggest that older adults may exhibit impaired glucose tolerance compared to younger populations [13, 14]. However, many studies focusing on insulin sensitivity (IS) and glucose parameters address the potential bias from age-related changes in body composition just by adjusting data for adiposity, which is less robust approach than a direct comparison of groups matched for this parameter. In addition, the analysis of FAHFAs remains challenging due to their low biological abundance. Consequently, most published studies focus only on selected FAHFA species or families, failing to capture the full complexity of FAHFA regulation.

Therefore, our aim was to analyze the effect of adiposity and age on FAHFA levels and composition in AT and circulation, and, in particular, to analyze the dynamics of circulating FAHFA levels in response to acute exercise. We used state-of-the-art targeted lipidomics to analyze AT and blood samples from women with a carefully assessed metabolic and cardiorespiratory fitness phenotype. We also analyzed the impact of exercise-mimicking conditions on FAHFA levels in adipocytes.

The results of this work point to a strong impact of obesity and aging on FAHFA levels in AT but not in circulation. Our findings also challenge the previously observed downregulation of FAHFA levels in response to acute exercise, and the presumption that SAT is the major source of circulating FAHFAs. On the other hand, the study highlights the importance of active lipolysis combined with increased free fatty acid (FFA) metabolism for FAHFA synthesis in adipocytes.

## 2. Methods and Subjects

Detailed description of methods as well as information on resources is provided in Supplementary Information and Tables.

### 2.1. Subjects and Study Design

The ETAPA clinical trial (NCT05572905) was approved by the Ethical Committee of the Third Faculty of Medicine, Charles University in Prague and the Ethical Committee of Kralovske Vinohrady University Hospital (EK-VP/40/0/2020), and conducted in accordance with the Declaration of Helsinki. Schemes of study flow and design are shown in Figure 1 A, B. Sixty Caucasian women were enrolled and stratified into groups (n=20) based on BMI (lean vs. obese) and age (young vs. older). To minimize confounding factors, lean and obese groups were age-matched, while young and older groups were individually matched for fat mass percentage (%FM) assessed by Dual-Energy X-ray Absorptiometry (DXA). Detailed inclusion/exclusion criteria are provided in the Supplementary Information.

**Figure 1.**
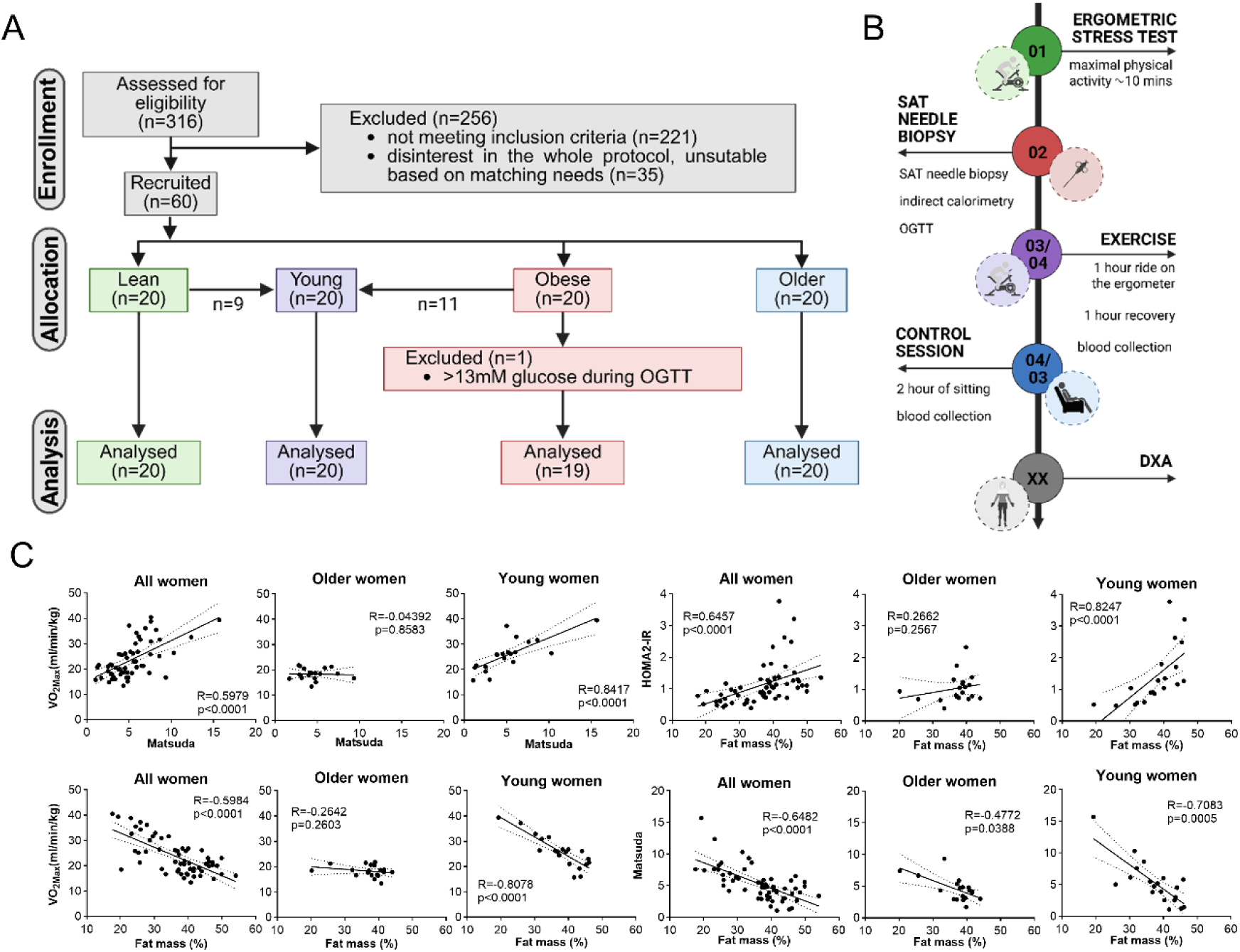
Experimental workflow and metabolic characteristics of the study cohorts. **A.** Clinical trial flow diagram**. B.** Clinical trial design diagram. **C**. Spearman correlations of selected parameters of adiposity, insulin sensitivity and cardiorespiratory fitness in all (n=59), young (n=20) and older women (n=20).

### 2.2. Baseline clinical investigations and AT sampling

Baseline assessments included body composition, VO_2max_ testing, respiratory quotients and resting metabolic rate. Glucose tolerance and various IS indexes were assessed by an oral glucose tolerance test (OGTT). Needle biopsies of superficial SAT were performed in the abdominal area under local anesthesia after an overnight fast.

### 2.3. Exercise and Control intervention

In a cross-over design, participants underwent two 2-hour interventions: 1) Acute exercise (EXE): 60 minutes of cycling at an intensity corresponding to maximal fat oxidation followed by 60 minutes of recovery; 2) Control (CON): 2 hours of seated rest. The order of the interventions was randomly assigned, both interventions were separated from each other by at least 1 week. Serial blood sampling was performed at 0, 60, and 120 minutes for FAHFA and metabolite analyses.

### 2.4. Plasma/serum analyses and calculations

Blood insulin and metabolites were analyzed in certified laboratories. Glycerol and FFAs were measured using colorimetric assays, serum protein levels were analyzed using ELISA. IS indexes were calculated as summarized by Gastaldelli [15] and using HOMA2 Calculator.

### 2.5. FAHFA and FFA analysis

Samples of AT (100 mg), plasma (100 μL), and *in vitro* cultivated adipocytes (grown in 2 wells of 6 well plate) were extracted according to previously described methods, with several modifications to improve extraction yield and FAHFA detection during LC-MS analysis [1, 16]. Samples were homogenized using a ball mill in a mixture of citric acid buffer and ethyl acetate (ratio 1:2, v/v) containing 1 ng of [^13^C_4_]-9-PAHSA as an internal standard. The ethyl acetate phase was collected and dried using a Savant SpeedVac. Based on optimization of extraction yield, we dissolved the dried extracts in 300 μL (200 μL for plasma) of chloroform and loaded the samples onto HyperSep SPE columns (500 mg/10 mL, 40–60 μm, 70 Å) for further purification by solid-phase extraction. FAHFAs were eluted with 5 mL ethyl acetate and dried. Before analysis, purified extracts were dissolved in 50 μL of a methanol/water mixture (95:5, v/v) to eliminate triglycerides, potentially suppressing FAHFA signal. Samples were analyzed by LC-MS as before [16]. FFAs in adipocytes were analyzed by untargeted LC/MS using a Vanquish UHPLC System coupled to an Orbitrap Exploris 480 mass spectrometer (Thermo Fisher Scientific, Bremen, Germany). FAHFA extracts were used, as the extraction procedure had been previously validated to fully retain FFA. Data were processed using MS-DIAL software (v4.9.221218) [17], and normalization was performed using locally estimated scatterplot smoothing (LOESS).

### 2.6. Culture of adipocytes

Isolation, culture and differentiation of human preadipocytes was described previously [18]. Cells were differentiated for 12 days and treated in serum-free medium (thus excluding the impact of exogenous FAHFA) for 48 hours with either 1 µM dexamethasone (DEX) or 250 µM 3-isobutyl-1-methylxanthine (IBMX) and harvested for the gene expression and FAHFA analysis. In the second experimental set, cells were exposed to 50 µM etomoxir (ETO), 10 µM thioridazine hydrochloride (TRD), or 250 µM IBMX, either alone or in combination (IBMX with ETO or TRD) and harvested for FFA and FAHFA analysis. Both experimental sets included five independent experiments.

### 2.7. Gene expression analysis

For mRNA analysis, total RNA was isolated and reverse transcribed as described previously [15]. For microfluidics, cDNA was preamplified and analyzed on Biomark Real Time qPCR system using 96.96 dynamic array. To analyze acyl-CoA synthetase long chain family member 1 (*ACSL1)* mRNA, qPCR was performed on Applied Biosystem 7500 Fast instrument. Data were normalized to the reference gene and analyzed after 2^−ΔCt^ or 2^−ΔΔCt^ calculation.

For protein analysis, cellular lysates in RIPA buffer were analyzed by Western blotting coupled with the ECL detection system.

### 2.8. Statistical analysis

Data normality was assessed and non-normal variables (including FAHFA levels in SAT and plasma) were log-transformed. Data are described as mean ± standard deviation or standard error of mean as indicated. Differences between groups, the impact of interventions and correlations were analyzed using two-way repeated measures ANOVA with (Tukey’s post/hoc), Multiple t-tests, Spearman’s and Pearson’s method, as appropriate. Significance was set at *p* < 0.05 and *q* < 0.05. All statistical analyses were performed using GraphPad Prism except for partial least squares-discriminant analysis (PLS-DA) and variable importance in projection (VIP) using MetaboAnalyst 6.0 [19].

## 3. Results

### 3.1. Baseline comparison of anthropometric, biochemical, and cardiometabolic parameters of the groups

The main characteristics of the subjects are described in Table 1. By design, adiposity parameters differed significantly between lean and obese, but were comparable between young and older women. The lean women had better metabolic health indices (e.g. glucose, insulin, HOMA2-IR, Matsuda, AdipoIR) and cardiorespiratory fitness (VO_2max_ or VO_2max_/FFM kg) than the obese (Table 1 and 2). Glucose tolerance (glucose AUC_OGTT_) was preserved in obese women through compensatory hyperinsulinemia. Basal FFA levels were also similar between groups despite higher AdipoIR in obese women.

**Table 1:** Clinical, biochemical, and insulin sensitivity characteristics of study groups. Data are expressed as mean ± SD. Data in italics were log transformed for statistical analysis based on a battery of normality tests. Multiple t-test with Welsch correction and Two stage step-up method of Benjamini, Krieger and Yekutieli post hoc analysis (*p* value, < 0.05; *q* value < 0.05) was used. HDL, high density lipoproteins; TAG triglycerides. AdipoIR, Adipose tissue Insulin Resistance index; AUC, area under the curve; iAUC, incremental area under the curve; BMI, body mass index; FFA, free fatty acids; HIRI, hepatic insulin resistance index; HOMA-IR, Homeostatic Model Assessment 2 – Insulin resistance; HOMA2-%B, Homeostatic Model Assessment 2 - Beta-cell function; IGI, Insulinogenic Index; OGTT, oral glucose tolerance test.

| Variable | Lean | Obese | Statistics |  | Young | Older | Statistics |  |
| --- | --- | --- | --- | --- | --- | --- | --- | --- |
|  | n = 20 | n = 19 | *p* value | *q* value | n = 20 | n = 20 | *p* value | *q* value |
| Age (years) | 29.6 ± 4.7 | 31.1 ± 3.9 | 0.3000 | 0.0944 | 29.7 ± 3.9 | 72.2 ± 3.8 | **<0.0001** | **<0.0001** |
| Weight (kg) | 60.0 ± 5.8 | 94.3 ± 9.5 | **<0.0001** | **<0.0001** | 79.2 ± 18.6 | 68.7 ± 8.9 | **0.0305** | 0.0958 |
| BMI (kg/m<sup>2</sup>) | 21.5 ± 1.6 | 34.1 ± 3.1 | **<0.0001** | **<0.00001** | 28.5 ± 6.4 | 26.4 ± 2.8 | 0.1776 | 0.2398 |
| Waist (cm) | 68.2 ± 3.5 | 93.4 ± 9.2 | **<0.0001** | **<0.0001** | 80.2 ± 13.5 | 82.8 ± 6.8 | 0.4551 | 0.4778 |
| Hips (cm) | 95.1 ± 6.1 | 120.5 ± 8.4 | **<0.0001** | **<0.0001** | 109.0 ± 13.7 | 101.6 ± 6.1 | **0.0406** | 0.0958 |
| Fat Mass (%) - DXA | 28 ± 5.5 | 45.2 ± 4.1 | **<0.0001** | **<0.0001** | 37.7 ± 7.1 | 37.3 ± 5.1 | 0.7664 | 0.7849 |
| Fat Free Mass (kg) - DXA | 43.3 ± 4.7 | 51.2 ± 3.5 | **<0.0001** | **<0.0001** | 47.3 ± 6.4 | 42.8 ± 4.5 | **0.0122** | 0.0576 |
| *TAG (mmol/L)* | *0.83 ± 0.21* | *1.15 ± 0.58* | **0.0324** | **0.0085** | *1.12 ± 0.57* | *1.15 ± 0.57* | *0.8305* | *0.7849* |
| HDL (mmol/L) | 1.69 ± 0.20 | 1.47 ± 0.34 | **0.0177** | **0.0053** | 1.56 ± 0.32 | 1.71 ± 0.42 | 0.2367 | 0.3195 |
| Total Cholesterol (mmol/L) | 4.65 ± 0.57 | 4.93 ± 0.91 | 0.2608 | 0.0608 | 4.86 ± 0.80 | 5.3 ± 0.79 | 0.1253 | 0.2369 |
| FFA (mmol/L) | 0.63 ± 0.30 | 0.59 ± 0.13 | 0.6153 | 0.1762 | 0.60 ± 0.20 | 0.71 ± 0.26 | 0.1321 | 0.2311 |
| *Glucose (mM)* | *4.94 ± 0.29* | *5.21 ± 0.35* | **0.0113** | **0.0035** | *5.04 ± 0.31* | *5.31 ± 0.30* | **0.0087** | **0.0127** |
| *Insulin (μIU/mL)* | *6.50 ± 1.87* | *14.8 ± 7.38* | **<0.0001** | **<0.0001** | *12.6 ± 8.57* | *9.03 ± 3.84* | *0.1889* | *0.1983* |
| *Glucose AUC<sub>OGTT</sub>* | *788 ± 135* | *831 ± 146* | *0.3446* | *0.099* | *806 ± 157* | *1005 ± 115* | **0.0001** | **0.0007** |
| *Glucose iAUC<sub>OGTT</sub>* | 216 ± 134 | 235 ± 149 | 0.8084 | 0.212 | 229 ± 160 | 384 ± 110 | **0.0059** | **0.0109** |
| *Insulin AUC<sub>OGTT</sub>* | 5967 ± 2110 | 13112 ± 9452 | **0.0004** | **0.0002** | 11600 ± 9259 | 9459 ± 4306 | 0.6451 | 0.4741 |
| *Insulin iAUC<sub>OGTT</sub>* | 5318 ± 2050 | 11725 ± 8891 | **0.0011** | **0.0005** | 10398 ± 8559 | 8633 ± 4188 | 0.7249 | 0.4844 |
| *HOMA-IR2* | 0.73 ± 0.21 | 1.65 ± 0.80 | **<0.0001** | **<0.0001** | 1.41 ± 0.94 | 1.03 ± 0.44 | 0.2255 | 0.2071 |
| *HOMA2%B* | 78.5 ± 14.7 | 122.0 ± 39.4 | **0.0001** | **<0.0001** | 112.0 ± 44.2 | 83.8 ± 21.8 | **0.0151** | **0.0185** |
| *Matsuda IS* | 7.42 ± 2.84 | 3.81 ± 1.65 | **<0.0001** | **<0.0001** | 5.23 ± 3.45 | 4.51 ± 1.79 | 0.9519 | 0.5382 |
| *HIRI* | 29.0 ± 22.0 | 88.5 ± 90.4 | **0.0029** | **0.0010** | 69.5 ± 70.2 | 48.4 ± 40.9 | 0.5201 | 0.4248 |
| *IGI* | 1.38 ± 0.95 | 3.13 ± 2.41 | **0.0029** | **0.0010** | 2.45 ± 1.74 | 1.02 ± 0.53 | **0.0003** | **0.0012** |
| *Disposition index* | 11.0 ± 12.5 | 11.2 ± 10.7 | 0.9981 | 0.2419 | 12.3 ± 13.7 | 4.41 ± 2.57 | **0.0017** | **0.0043** |
| *AdipoIR* | 4.28 ± 2.74 | 8.41 ± 3.85 | **<0.0001** | **<0.0001** | 7.47 ± 4.65 | 6.51 ± 3.66 | 0.8500 | 0.5206 |

**Table 2:** Indirect calorimetry and fitness characteristics of study groups in basal state, response to OGTT, and during EXE intervention. Data in italics were log transformed for statistical analysis based on a battery of normality tests, Multiple t-test with Welsch correction and Two stage step-up method of Benjamini, Krieger and Yekutieli post hoc analysis (*p* value, < 0.05; *q* value < 0.05) was used. EXE, acute exercise intervention; FFM, fat free mass; P, power; RMR, resting metabolic rate; RQ, respiratory quotient.

| Variable | Lean | Obese | Statistics |  | Young | Older | Statistics |  |
| --- | --- | --- | --- | --- | --- | --- | --- | --- |
|  | n = 20 | n = 19 | p value | q value | n = 20 | n = 20 | p value | q value |
| RMR | 1370 ± 128 | 1744 ± 189 | **<0.0001** | **<0.0001** | 1542 ± 260 | 1320 ± 145 | **0.0029** | **0.0030** |
| RMR/FFM | 31.9 ± 3.7 | 34.3 ± 3.6 | **0.0484** | 0.0678 | 32.8 ± 3.7 | 31.1 ± 3.2 | 0.0964 | 0.0868 |
| RQ<sub>basal</sub> | 0.75 ± 0.06 | 0.74 ± 0.05 | 0.4381 | 0.3944 | 0.75 ± 0.06 | 0.74 ± 0.05 | 0.7826 | 0.4109 |
| RQ<sub>OGTT120</sub> | 0.84 ± 0.05 | 0.79 ± 0.05 | **0.0038** | **0.0080** | 0.82 ± 0.06 | 0.85 ± 0.06 | 0.2569 | 0.1619 |
| Relative Δ RQ<sub>basal</sub>-OGTT120 (%) | 12.47 ± 7.29 | 7.54 ± 7.04 | **0.0411** | 0.0678 | 10.27 ± 8.04 | 14.09 ± 7.73 | 0.1387 | 0.1093 |
| $P_{max}$ (W) | 174.5 ± 41.8 | 176.5 ± 37.2 | 0.8140 | 0.5698 | 178.4 ± 37.7 | 105.2 ± 21.3 | **<0.0001** | **<0.0001** |
| VO<sub>2max</sub> (ml/min/kg) | 31.5 ± 5.3 | 20.8 ± 3.8 | **<0.0001** | **<0.0001** | 25.9 ± 6.3 | 18.3 ± 2.2 | **<0.0001** | **<0.0001** |
| VO<sub>2max</sub>FFM (ml/min/kg FFM) | 43.5 ± 5.6 | 37.7 ± 5.7 | **0.0028** | **0.0079** | 41.3 ± 5.6 | 29.7 ± 3.2 | **<0.0001** | **<0.0001** |
| Heart rate<sub>max</sub> | 177.8 ± 10.4 | 174.4 ± 9.5 | 0.3237 | 0.3399 | 178.9 ± 6.9 | 137.9 ± 18.0 | **<0.0001** | **<0.0001** |
| % Heart rate<sub>max</sub> during EXE | 67.40 ± 5.90 | 66.51 ± 4.80 | 0.6199 | 0.4734 | 67.75 ± 5.49 | 71.28 ± 10.76 | 0.2285 | 0.1600 |
| % VO<sub>2max</sub> during EXE | 48.30 ± 4.39 | 47.06 ± 5.65 | 0.4696 | 0.3944 | 48.81 ± 3.78 | 57.69 ± 9.45 | **0.0007** | **0.0008** |
| % P<sub>max</sub> during EXE | 27.57 ± 3.40 | 25.99 ± 4.92 | 0.2696 | 0.3236 | 27.11 ± 4.92 | 25.96 ± 3.71 | 0.4285 | 0.2454 |

Older and young women showed similar lipid profiles and IS indices (HOMA2-IR, AdipoIR, HIRI, Matsuda). However, older women had higher fasting glucose and glucose AUC_OGTT_ associated with diminished pancreatic β-cell secretory capacity (HOMA2-%B, IGI, Disposition index), and lower cardiorespiratory fitness (Heart rate_max_, VO_2max_ and VO_2max/FFM kg_). Metabolic flexibility calculated as relative delta RQ_basal-OGTT120_ tended to be higher in lean vs. obese, but was similar between young and older.

When the parameters from all women were correlated, adiposity correlated negatively with VO_2max_ and IS, but these associations were attenuated or absent in older women (Figure 1C).

### 3.2. FAHFA levels in subcutaneous adipose tissue

In SAT, we have detected 36 FAHFA regioisomers, with total FAHFA levels ranging from 25.6 to 324.8 pmol/g (mean 97.9 pmol/g). We compared the absolute levels as well as the molar ratio of these regioisomers and their families defined by their fatty acid composition. Both adiposity and age affected the majority of detected FAHFAs (24 and 23 regioisomers, respectively). Specifically, 15 of these regioisomers, total FAHFA levels (mean ± SEM, lean: 109.3 ± 16.6 vs obese: 64.1 ± 4.9 pmol/g; young: 82.6 ± 10.9 vs. older: 118.8 ± 14.7 pmol/g), and the PAHSA, PAHOA, OAHOA, and LAHOA families were higher in lean and older women compared to their counterparts (Figure 2A and B). To investigate this unexpected age-related increase in SAT FAHFA content (despite generally higher adiposity), we sub-analyzed fat mass-matched lean young and older women (n= 9, FM %, lean young: 31.6 ± 5.8; older: 33.1 ± 6.2). This confirmed that age increased SAHSA levels above those in lean subjects (mean ± SEM, lean young: 4.2 ± 0.5; lean older: 10.5 ± 2.0 pmol/g, *p* = 0.0002, *q* = 0.0021).

**Figure 2.**
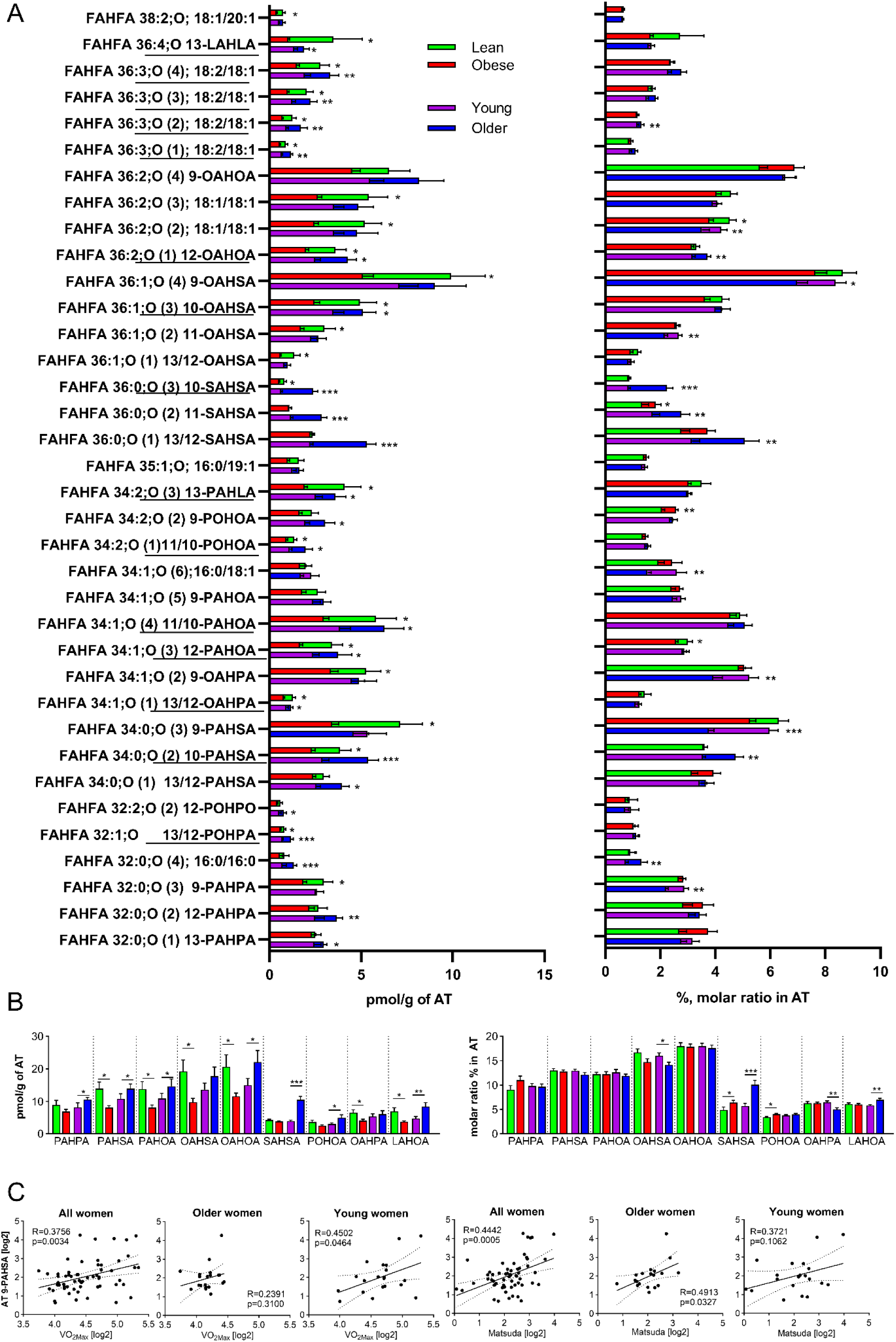
FAHFA levels in SAT. **A.** Absolute and relative levels of FAHFA regioisomers in SAT**. B** Absolute and relative levels of FAHFA families in SAT. Data are expressed as mean ± SEM; lean-green, obese- red, young- violet, older- blue, Multiple-t tests of log transformed data, difference between the groups, * *q* < 0.05, ** *q* < 0.01, *** *q* < 0.001. **C.** Pearson correlations of log transformed baseline AT 9-PAHSA level with VO_2max_ and Matsuda index in all (n=59), young (n=20) and older women (n=20). LAHLA, linoleic acid ester of hydroxy-linoleic acid; LAHOA, linoleic acid ester of hydroxy oleic acid; OAHOA, oleic acid ester of hydroxy-oleic acid; OAHPA, oleic acid ester of hydroxy-palmitic acid; OAHSA, oleic acid ester of hydroxy-stearic acid; PAHLA, palmitic acid ester of hydroxy-linoleic acid; PAHOA, palmitic acid ester of hydroxy-oleic acid; PAHPA, palmitic acid ester of hydroxy-palmitic acid; PAHSA, palmitic acid ester of hydroxy-stearic acid; POHOA, palmitoleic acid ester of hydroxy-oleic acid; POHPA, palmitoleic acid ester of hydroxy-palmitic acid; POHPO, palmitoleic acid ester of hydroxy-palmitoleic acid; SAHSA, stearic acid ester of hydroxy-stearic acid.

Moreover, adiposity and age had a differing impact on the position of branching carbon: adiposity primarily altered 9-carbon branched isomers (e.g. 9-PAHSA, 9-OAHSA), whereas age affected mostly saturated 13/12-carbon branched species (e.g. 13/12-PAHSA, 13/12-SAHSA). Still, the levels of insulin-sensitizing 9-PAHSA were preserved in lean aged SAT but reduced by obesity (mean ± SEM, lean young: 6.8 ± 2.1 pmol/g vs lean older: 6.1 ± 1.7 pmol/g, *p*= 0.9821; obese young: 3.7 ± 0.5 pmol/g vs obese older: 3.4 ± 0.3 pmol/g, *p*= 0.8384).

According to molar ratios, 9-OAHSA was the most abundant regioisomer (7.0-8.6%) and OAHOA the most abundant family (17.6-18.0% of total FAHFAs) across all groups in SAT (Figure 2A and B). We found adiposity had minimal impact on relative composition of FAHFA, when only the SAHSA family differed between the groups by more than 1%. In contrast, aging significantly altered the molar ratios of 14 regioisomers (eight of them differing by more than 1%) and nearly doubled the SAHSA family proportion (5.7% to 10.1%) in older compared to young women. Interestingly, within the PAHSA and OAHOA families, the regioisomers differing in the nearest branching point (i.e. 9- and 10-PAHSA) were affected by age in opposite directions. In other words, aging shifted the branching point toward carbons more distant from the carboxyl end. Thus, while adiposity affected mainly absolute levels of FAHFA, aging altered both the absolute and relative content of FAHFA, particularly SAHSA, as well as the position of the branching carbon.

To assess the relationship to cardiometabolic health, we correlated FAHFA SAT levels with VO_2max_ and IS indices (Figure 2C, Supplementary Table S2). In the total cohort, the Matsuda index correlated with both 9-PAHSA and total FAHFA, while VO_2max_ correlated with 9-PAHSA only. In older women, the association of 9-PAHSA with IS was preserved but the correlation between 9-PAHSA and VO_2max_ was lost.

### 3.3. FAHFA levels in plasma

In plasma, we have detected 63 FAHFA regioisomers, i.e. almost twice as many as in SAT. Overall circulating FAHFA levels ranged from 19.29 to 66.72 pmol/mL, with the mean 32.36 pmol/mL. Day-to-day variability appeared to be about 35%, similar to FFA and insulin (data not shown). According to molar ratios, the most abundant regioisomer in plasma irrespectively of the group was LAHOA 36:3;O (FAHFA18:2/18:1) and the most abundant family was PAHOA (Figure 3A, Supplementary table S3). These relative abundancies, which are not biased by the effect of matrix during the extraction procedure, show that the most abundant FAHFA regioisomers and families in plasma do not correspond to those found in SAT.

**Figure 3.**
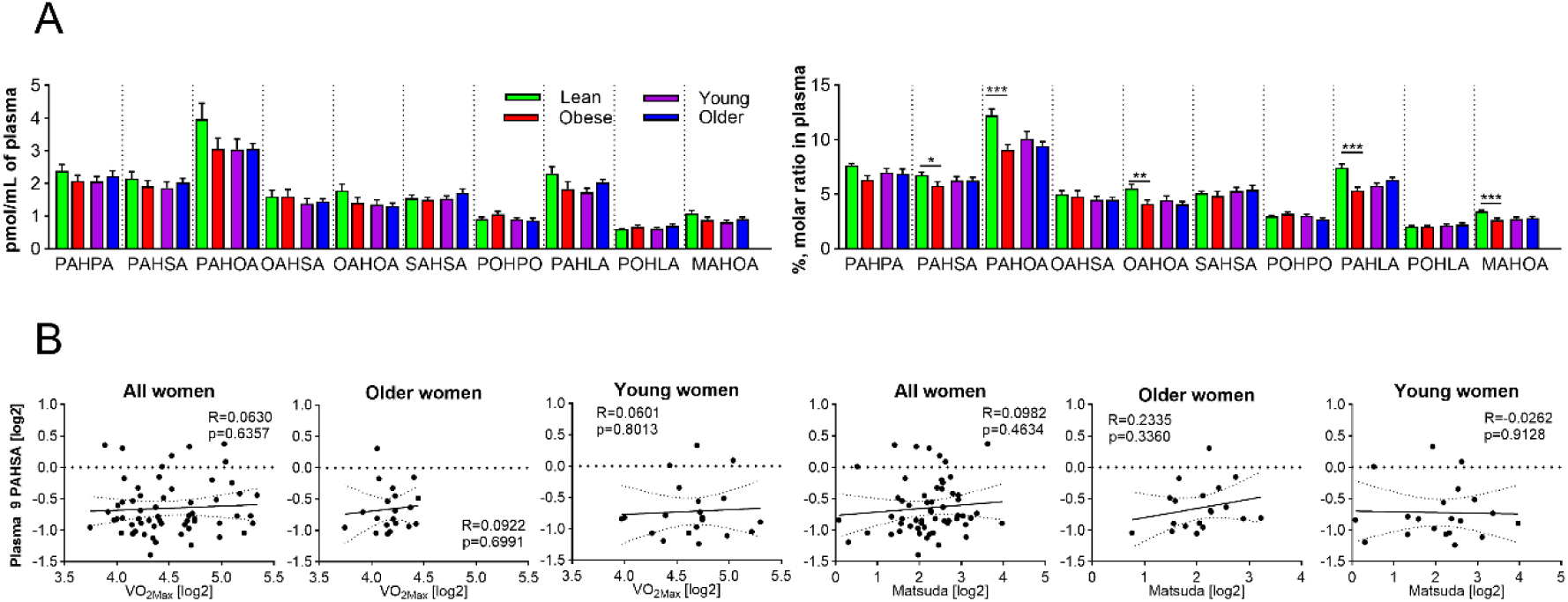
FAHFA levels in circulation. **A.** Absolute and relative levels of FAHFA families in circulation. Data are expressed as mean ± SEM; lean-green, obese- red, young- violet, older- blue, Multiple-t tests of log transformed data, difference between the groups, * *q* < 0.05, ** *q* < 0.01, *** *q* < 0.001. **C.** Pearson correlations of log transformed baseline circulating 9-PAHSA level with VO_2max_ and Matsuda index in all (n=59), young (n=20) and older women (n=20). MAHOA, myristic acid ester of hydroxy-oleic acid; OAHOA, oleic acid ester of hydroxy-oleic acid; OAHSA, oleic acid ester of hydroxy-stearic acid; PAHLA, palmitic acid ester of hydroxy-linoleic acid; PAHOA, palmitic acid ester of hydroxy-oleic acid; PAHPA, palmitic acid ester of hydroxy-palmitic acid; PAHSA, palmitic acid ester of hydroxy-stearic acid; POHLA, palmitoleic acid ester of hydroxy-linoleic acid; POHPO, palmitoleic acid ester of hydroxy-palmitoleic acid; SAHSA, stearic acid ester of hydroxy-stearic acid.

In contrast to SAT, total circulating FAHFA levels were similar across all the groups. Only three FAHFA regioisomers were increased by adiposity (FAHFA 17:1/18:1; FAHFA 20:4/16:0; FAHFA 18:1/20:1) and one (FAHFA 18:1/19:1) by age (Supplementary Table S3). While age had no impact on molar ratios, adiposity significantly altered the relative FAHFA composition (32 out of 63 regioisomers). The most prominent differences included a four-fold increase in the molar ratio of 13/12-AAHPA (FAHFA 20:4/16:0) and a 3% reduction in the PAHOA family in obese vs. lean women (Figure 3A, Supplementary Table S3). Together, adiposity rather than age affected relative levels of circulating FAHFAs despite having no effect on total FAHFA content, which differs from what we observed in SAT.

To determine the relationship between adiposity, cardiometabolic fitness, IS and FAHFAs (especially 9-PAHSA), a correlation analysis was performed using the entire cohort (n=59), and also separately for groups of older and young women. However, there was no positive correlation between either 9-PAHSA or any other FAHFA levels and the parameters mentioned, in any of the groups (Figure 3B, Supplementary Table S2). Furthermore, while strong correlations existed among regioisomers within SAT, only weak and isolated correlations were found between SAT and plasma FAHFA levels (Supplementary Figure S1), further questioning SAT as the primary source of circulating FAHFAs.

### 3.4. Impact of exercise on FAHFA and cytokines in the circulation

To evaluate FAHFA kinetics in response to an acute bout of exercise, participants completed two interventions: 60 minutes of exercise at a constant intensity followed by 60 minutes of rest (EXE), and a control intervention of the same duration without physical activity (CON), to account for the impact of fasting and circadian rhythms. The EXE intervention was performed at similar levels of % P_max_ and % Heart rate_max_ across all groups (Table 2). The EXE intervention induced lipolysis in all groups, evidenced by increased circulating FFA and glycerol levels, whereas no such changes occurred during CON (Figure 4A and B). Thus, EXE intervention clearly enhanced lipid mobilization beyond the effects of fasting.

**Figure 4.**
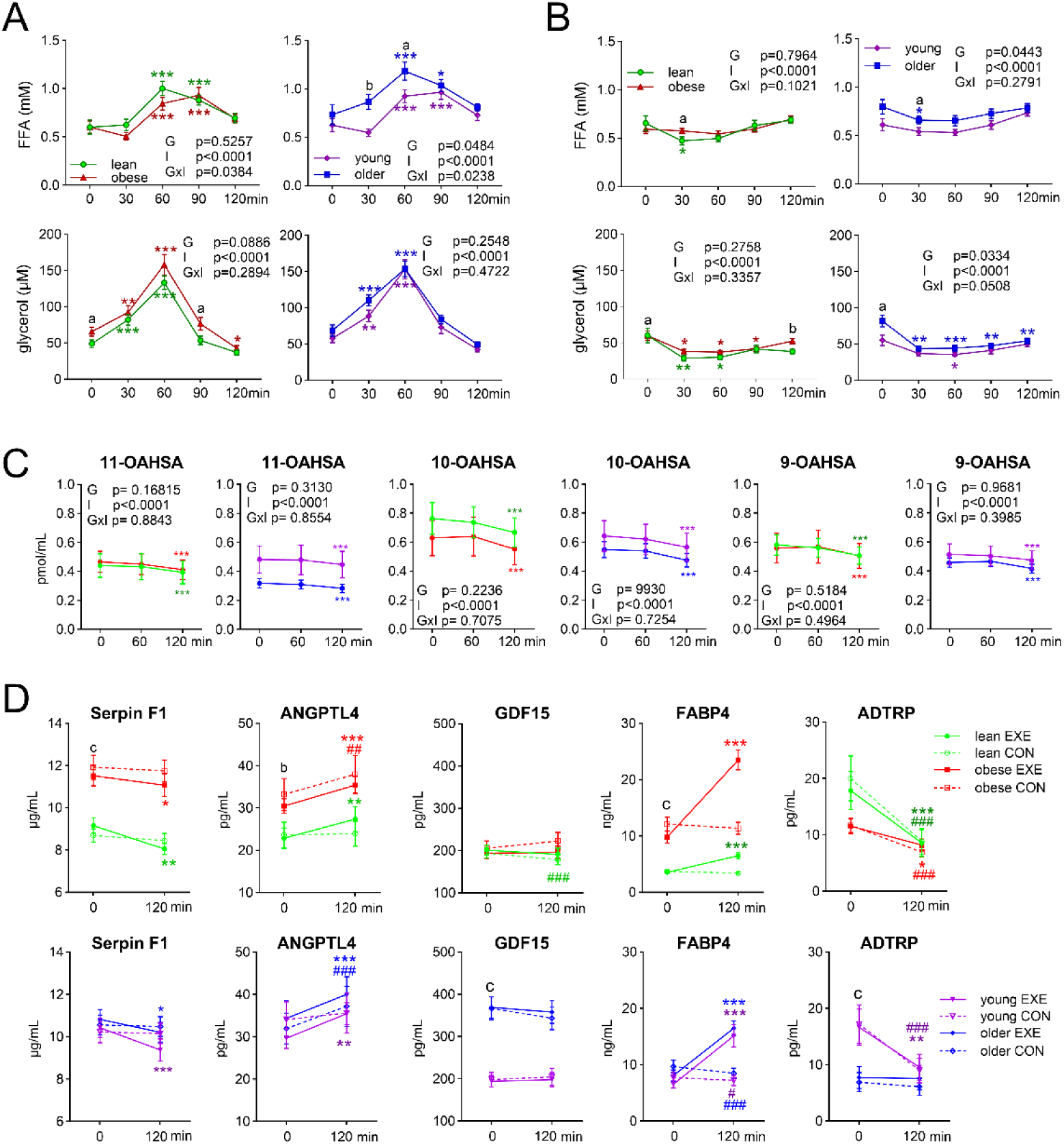
Blood profiles of metabolites and cytokines during the interventions. **A.** Blood FFA and glycerol during EXE intervention. **B**. Blood FFA and glycerol during CON intervention. **C.** Plasma level of regiosiomers from the OAHSA family in response to EXE. **D.** Serum levels of cytokines in response to EXE and CON interventions. Data are expressed as mean ± SEM; lean-green, obese- red, young- violet, older- blue. Statistics for **A, B, C**: Two-way ANOVA, I-effect of intervention, G-effect of group, GxI-interaction between the factors; posthoc testing: difference from baseline, \**p* < 0.05, ** *p* < 0.01, *** *p* < 0.001; difference between the groups, ^a^ < 0.05, ^b^ < 0.01, ^C^ < 0.001. Statistics for **D**: Two-way ANOVA, I-effect of intervention, G-effect of group, GxI-interaction between the factors: Serpin F1 lean vs. obese, I *p* < 0.0001, G *p* < 0.0001, GxI *p* = 0.0055; young vs. older, I *p* < 0.0001, G *p* = 0.4066, GxI *p* = 0.1043; ANGPTL4 lean vs. obese, I *p* = 0.0084, G *p* = 0.0036, GxI *p* = 0.2234; young vs. older, I *p* = 0.0005, G *p* = 0.6593, GxI *p* = 0.3151; GDF15 lean vs. obese, I *p* = 0.5121, G *p* = 0.5594, GxI *p* < 0.0001; young vs. older, I *p* = 0.7198, G *p* < 0.0001, GxI *p* = 0.3975; FABP4 lean vs. obese, I *p* < 0.0001, G *p* < 0.0001, GxI *p* = 0.0133; young vs. older, I *p* < .0001, G *p* = 0.1100, GxI *p* = .8728; ADTRP lean vs. obese, I *p* < 0.0001, G *p* = 0.5342, GxI *p* = 0.0184; young vs. older, I *p* < 0.0001, G *p* = 0.0028, GxI *p* = 0.0009; posthoc testing: impact of EXE intervention * *p* < 0.05, ** *p* < 0.01, *** *p* < 0.001; impact of CON intervention ^#^ *p* < 0.05, ^##^ *p* < 0.01, ^###^ *p* < 0.001 difference between the groups ^a^ *p* < 0.05, ^b^ *p* < 0.01, ^c^ *p* < 0.001. FFA, free fatty acids; CON, control intervention; EXE, acute exercise intervention. ADTRP; Androgen Dependent TFPI Regulating Protein; ANGPTL4, Angiopoietin-like protein 4; FABP4, Fatty Acid Binding Protein 4; GDF15, Growth Differentiation Factor 15; OAHSA, oleic acid ester of hydroxy-stearic acid.

We then analyzed plasma levels of FAHFA regioisomers and families at the beginning and end of both interventions. Within the EXE session, we observed a decrease in 53 species in lean and 21 in older women, particularly within the OAHSA family (Figure 4C, Supplementary Table S4), whereas only a few regioisomers were affected in the obese and young women. This decline occurred during the recovery period (120 min) rather than immediately post-exercise (60 min). However, when comparing the absolute and relative changes between the two interventions (delta 0–120 min, CON vs. EXE), no EXE-specific effects were detected (Supplementary Table S5). Thus, acute exercise does not appear to specifically modulate circulating FAHFAs beyond the effects of fasting and circadian rhythms.

We further analyzed circulating levels of cytokines that have been previously shown to react to acute exercise or to be involved in the regulation of lipolysis, IS, and FAHFA turnover [20–24]. Baseline levels of Angiopoietin-like protein 4 (ANGPTL4), Fatty Acid Binding Protein 4 (FABP4) and serpin F1 were higher in obese vs. lean women but were independent of age, while Growth Differentiation Factor 15 (GDF15) and Androgen Dependent TFPI Regulating Protein (ADTRP) levels were affected by age (GDF15 being higher and ADTRP lower in older women), but not by adiposity (Supplementary Table S5).

In response to EXE, Serpin F1 decreased and FABP4 increased across all groups, with the Serpin F1 response being more pronounced in lean and FABP4 in obese women (Figure 4D). EXE specifically increased ANGPTL4 only in lean and young women, while ADTRP levels decreased during both EXE and CON in all younger groups. Notably, baseline circulating levels of ADTRP, the putative FAHFA hydrolase, did not correlate with circulating FAHFA levels (Supplementary Table S2). GDF15 levels were not affected by EXE in any group. Together, cytokines mostly affected by adiposity appeared to be more responsive to EXE intervention.

### 3.5. FAHFA levels in adipocytes *in vitro*

To investigate how stimuli related to acute exercise—specifically lipolysis and glucocorticoid signaling [25, 26]—affect FAHFA levels, human *in vitro* differentiated adipocytes were treated with either 250 µM IBMX (lipolytic stimulant) or 1 µM DEX (synthetic glucocorticoid) for 48 hours in serum-free medium.

At the mRNA level, both DEX and IBMX altered most of the analyzed genes (85 and 53 out of 94, respectively), which were selected based on their potential involvement in the regulation of FAHFA, lipid, and glucose metabolism, mitochondrial and peroxisome biogenesis/function, inflammation, oxidative stress and other processes (Supplementary Table S1, S6). Notably, 24 genes were regulated similarly by both treatments, including the upregulation of peroxiredoxin 6 (*PRDX6),* patatin like domain 2, triacylglycerol lipase *(PNPLA2)*, and lipase E (*LIPE)*, i.e. genes involved in FAHFA synthesis and turnover, and the downregulation of the FAHFA hydrolase *ADTRP*, suggesting a shift toward increased FAHFA synthesis and stability.

However, only IBMX treatment increased levels of 39 regioisomers out of 60 detected FAHFAs, 7 FAHFA families (Figure 5A, Supplementary Table S6), and also total FAHFA levels (mean ± SEM; control: 1, DEX: 1.16 ± 0.05, IBMX: 1.62 ± 0.10). To elucidate the mechanisms behind this specific response, we combined several approaches to identify the metabolic pathway potentially responsible for this IBMX-driven increase of intracellular FAHFA levels. First, we focused on 25 genes that were inversely regulated by DEX and IBMX, i.e., upregulated in response to IBMX and downregulated in response to DEX. These genes included genes important for lipid metabolism (e.g. *FABP4;* peroxisome proliferator-activated receptorγ, *PPARγ*; diacylglycerol O-acyltransferase 2*, DGAT2;* adrenergic receptor β1, *ADRB1*) as well as those important for mitochondrial and peroxisomal fatty acid oxidation (FAO) (acetyl-CoA acyltransferase 1, *ACAA1;* carnitine palmitoyltransferase 1B, *CPT1B;* enoyl-CoA hydratase/3-hydroxyacyl CoA dehydrogenase, *EHHADH*, hydroxyacyl-CoA dehydrogenase trifunctional multienzyme complex subunit α, *HADHA;* succinate dehydrogenase complex flavoprotein subunit α, *SDHA*), and for oxidative stress response (e.g. catalase, *CAT*; heme oxygenase 1, *HO1*). We then employed PLS-DA analysis in combination with VIP scores to identify the key genes contributing to the separation of experimental conditions (Figure 5B). This approach identified both *PNPLA2 and LIPE* as being among the genes with the highest discriminatory power. Finally, correlation analysis revealed that the highest number of FAHFA regioisomers associated with several genes related to lipolysis and β-oxidation, e.g. peroxisome proliferator-activated receptor γ coactivator 1α *(PGC1α), ANGPTL4, LIPE* and *ACAA1* mRNA, whereas the expression of *PRDX6* and androgen induced 1 (*AIG1)* showed no substantial relationship with FAHFA levels (Figure 5C, Supplementary Table S6). Furthermore, protein analysis confirmed that IBMX increased both ATGL and activated HSL (Figure 5D). IBMX also upregulated mRNA expression of *ACSL1*, the enzyme, which promotes FFA re-esterification or utilization via β-oxidation (Figure 5E).

**Figure 5.**
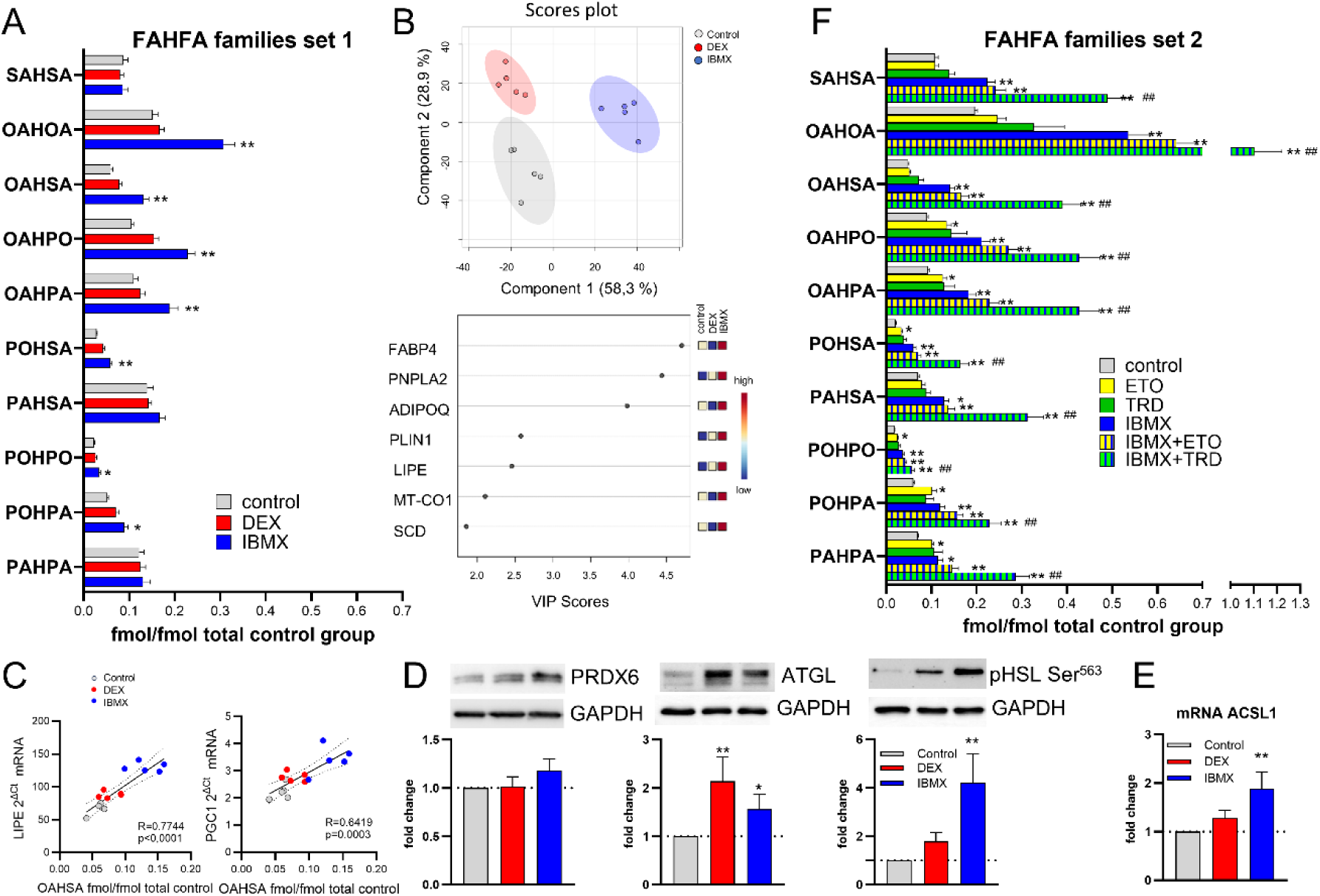
Response of *in vitro* human adipocytes to the exercise-mimicking conditions. Adipocytes differentiated *in vitro* were treated for 48 hrs with vehicle, Dexamethasone, IBMX, Etomoxir, Thioridazine or combinations of these drugs **A.** Levels of FAHFA in families normalized to total FAHFA content in control cells, set I. Data are expressed as mean ± SEM; Multiple-t tests, difference between the groups, * *q* < 0.05, ** *q* < 0.01, *** *q* < 0.001. **B.** PLS-DA of 94 analyzed genes and genes with VIP scores >1.5. **C.** Spearman correlations between mRNA levels of LIPE and PGC1 and levels of OAHSA family. **D**. Protein levels of PRDX6, ATGL, HSL p-Ser563 normalized to GAPDH levels. Mann Whitney t test, * *p* < 0.05, ** *p* < 0.01. Representative western blots are shown. **E**. mRNA levels of *ACSL1*, ** *p* < 0.01. **F.** Levels of FAHFA in families normalized to total FAHFA content in control cells, set II. Data are expressed as mean ± SEM; Multiple-t tests, difference compared to Control, * *q* < 0.05, ** *q* < 0.01, or to IBMX ^##^ *q* < 0.01. OAHOA, oleic acid ester of hydroxy-oleic acid; OAHPA, oleic acid ester of hydroxy-palmitic acid; OAHSA, oleic acid ester of hydroxy-stearic acid; PAHPA, palmitic acid ester of hydroxy-palmitic acid; PAHSA, palmitic acid ester of hydroxy-stearic acid; POHPA, palmitoleic acid ester of hydroxy-palmitic acid; POHPO, palmitoleic acid ester of hydroxy-palmitoleic acid; POHSA, palmitoleic acid ester of hydroxy-stearic acid; SAHSA, stearic acid ester of hydroxy-stearic acid; ACSL1, acyl-CoA synthetase long chain family member 1; PNPLA2, patatin like domain 2, triacylglycerol lipase (ATGL), adipose tissue triglyceride lipase; GAPDH, glyceraldehyde-3-phosphate dehydrogenase; LIPE, lipase E (HSL); PGC1α, peroxisome proliferator-activated receptor γ coactivator 1-α; PRDX6, peroxiredoxin 6; ETO, etomorix; TRD, thioridazine, DEX, dexamethasone; IBMX, 3-isobutyl-1-methylxanthine; VIP, Variable Importance in Projection.

Together, while an active lipolysis was necessary for *in vitro* FAHFA synthesis, our results suggested that increased FFA utilization might play a regulatory role. To test this, we inhibited mitochondrial and peroxisomal FAO using etomoxir (ETO) and thioridazine (TRD) [27], respectively, under both basal and IBMX-stimulated conditions. The efficacy of these inhibitors was confirmed by the significant accumulation of total as well as very-long-chain FFAs (Supplementary Table S6). Notably, contrary to our initial hypothesis, FAO inhibition did not reduce FAHFA synthesis; instead, mitochondrial FAO inhibition partially mimicked the IBMX effect (Figure 5F, Supplementary table S6). Although TRD treatment alone was insufficient to trigger FAHFA production, its combination with IBMX-induced lipolysis synergistically enhanced FAHFA levels, surpassing the effect of IBMX alone (Figure 5F, Supplementary table S6). Thus, FAHFA synthesis can be further augmented by suppression of peroxisomal FAO. Taken together, our results indicate a substrate-competing mechanism where FAHFA synthesis is favored when lipolytic supply is high and the degradative pathways for their precursors are restricted.

## 4. Discussion

### 4.1. Study design

The study design in untrained women allowed us to distinguish the independent impacts of adiposity and age on cardiometabolic parameters and FAHFA levels, while assessing the impact of acute exercise on circulating FAHFA levels. The adiposity-independent effect of age was ensured by comparing young and older women who were matched for FM percentage. Their similar waist circumferences further suggest comparable amounts of visceral AT, a key determinant of IS in older individuals [28]. Moreover, a parallel control intervention was included to account for potential confounders such as circadian rhythms or prolonged fasting.

### 4.2. FAHFA in SAT

We have shown that both adiposity and age differentially affect FAHFA levels, relative composition and branching patterns in SAT. While obesity was associated with lower levels of FAHFA branched at carbon 9 (9-C branched), advanced age was associated with higher levels the SAHSA family and FAHFAs branched at carbon 12 and above (e.g., 13,12-C branched). This finding has a potential biological significance as 13-C branched FAHFAs are less potent than 9-C branched FAHFAs in stimulating adipocyte glucose uptake and mitigating TNFα-induced inflammation [3].

Although our findings of higher SAHSA levels in SAT of older women mirror the accumulation of SAHSAs seen in the visceral AT of aged mice, we have not observed higher content of LAHLA, which may stem from the different type of fat depot studied [29].

In aged mice, the accumulation of FAHFA in visceral AT is attributed to the increased liberation of FAHFAs from their storage form, i.e., triglycerides, via HSL and ATGL-mediated lipolysis [29–31]. However, in our study, circulating FFA levels at baseline were similar between young and older women, suggesting that overall lipolytic activity was comparable. Instead, age-related reactive oxygen species (ROS) accumulation—driving lipid peroxidation and subsequently generating hydroxy-fatty acids (HFAs)—seems a more plausible mechanism for the increase in total FAHFAs as well as observed shift in branching, though the exact pathway remains unknown.

Finally, we noted considerable interindividual variability of FAHFA levels within groups. This may reflect the type of regular diet and saturated FFA consumption, which appear to affect FAHFA levels in humans [9].

### 4.3. Circulating FAHFA levels and their source

Regarding circulating FAHFA levels, we observed no substantial differences between the study groups. Our absolute circulating FAHFA levels align with previously published data [1, 11], although they are slightly higher than those reported by Kellerer et al. [9]. Notably, however, FAHFA levels in our study are over an order of magnitude lower than those reported by Nelson et al. [12], who also observed significant baseline variations (2- to 8-fold differences) between normal-weight and overweight individuals— differences not reflected in our data. It is challenging to explain such a discrepancy solely by differences in the study participants, though Nelson et al. used trained subjects, who can be expected to have higher FAHFA levels [11], and groups with an uneven sex distribution, despite documented sex-based differences in both, adiposity and FAHFA levels [32]. Nevertheless, we suspect that the primary cause of the large discrepancy in circulating FAHFA levels between our study and that of Nelson et al. relates to sample processing, specifically the chemical derivatization of FAHFA. Importantly, we found no correlation between circulating and SAT FAHFA levels, suggesting that SAT is not the primary source of circulating FAHFAs; instead, hepatic FAHFAs could be proposed as an alternative source. Indeed, even the original report by Yore et al. [1] described distinct behaviors in AT, liver, and circulating FAHFAs during fasting, which increased FAHFA levels in AT but not in the circulation and the liver. Furthermore, while we observed a positive correlation between SAT 9-PAHSA and IS, circulating 9-PAHSA levels showed no such association. Similarly, the correlation between IS and circulating 9-PAHSA demonstrated by Yore et al. [1] was considerably weaker than the correlation with SAT 9-PAHSA levels. Thus, we propose that FAHFAs released from AT act primarily within the AT microenvironment, supporting whole-body IS indirectly by preventing ectopic fat accumulation.

### 4.4. Exercise effect on circulating FAHFA levels

One of the key objectives of our study was to evaluate the effects of acute exercise on circulating FAHFA levels, a topic previously addressed only by Nelson et al. [12], who reported a 40–79% decrease in most FAHFA families in lean, but not overweight, recreational runners. [12]. Similarly, we observed a decline in FAHFA levels–particularly within the OAHSA family- in lean and older sedentary women, although this decrease was less pronounced. This could be related to the differences in fitness and adiposity level of participants included to the studies (Nelson et al.: FM ∼ 19%, VO_2max/FFMkg_ ∼ 73.3, 6 women/8 men; our study: FM ∼ 28%, VO_2max_/_FFMkg_ ∼ 43.5, 20 women).

Interestingly, OAHSA appears to be one of the most dynamic FAHFA families, sensitively reflecting various metabolic conditions. For instance, 9-OAHSA is strongly responsive to bariatric surgery-induced weight loss [9] and negatively correlates with glucose levels in humans [33]. Experimental models further support its metabolic importance: showing cardioprotective and anti-inflammatory effects, as well as a beneficial effect on mitochondrial function [33, 34]. However, our control intervention clearly demonstrated that the observed decline in OAHSA was not specific to physical activity itself. Moreover, this decline was unrelated to systemic lipolysis, which was not induced during the control session. Consequently, we hypothesize that circulating FAHFA levels may be subject to circadian regulation. Such a mechanism would also explain the diminished FAHFA dynamics observed in women with obesity, a condition frequently associated with circadian rhythm dysregulation [35], which extends to liver metabolism [36].

Additionally, circadian rhythms may also affect circulating levels of ADTRP, which decreased in both interventions but did not correlate with FAHFA levels. In fact, it remains unclear whether circulating ADTRP shed from its transmembrane form retains FAHFA hydrolase activity. In contrast, acute exercise robustly increased circulating FABP4 levels, consistent with a recent report [23]. Interestingly, the elevation of FABP4 was more pronounced in obese women. Given the established positive correlation between FABP4 and cardiovascular disease [37], this finding raises questions regarding the clinical implications of such an increase. However, it has been suggested that FABP4 may potentiate insulin secretion by β-cells, a role that appears to be more prominent in the context of obesity [38]. Finally, the exercise-induced decrease in Serpin F1, a negative regulator of muscle contraction [39], may represent a beneficial adaptation for muscle health in older individuals.

### 4.5. *In vitro* Regulation of FAHFA Synthesis

Our study on human adipocytes provides insights into the regulation of intracellular FAHFA levels. While both DEX and IBMX treatments increased lipolytic genes and *PRDX6*, only IBMX treatment significantly elevated FAHFA content. This indicates that gene induction by DEX is insufficient, or that these changes alone cannot drive FAHFA synthesis. Furthermore, the role of PRDX6 for FAHFA synthesis—originally identified in mice [40] —remains questionable in human adipocytes; its mRNA levels did not correlate with FAHFA levels, and our unpublished proteomic data from human adipocytes indicate that PRDX6 protein levels are constitutively high. Nevertheless, we confirmed that chronic lipolysis stimulation is linked to increased FAHFA synthesis, consistent with previous reports in 3T3-L1 adipocytes [30].

We also explored additional regulatory pathways. The IRF3-mediated upregulation of AIG1 [41] was excluded, as IBMX had no impact on *AIG1* expression, and DEX failed to increase FAHFA levels despite reducing *AIG1* mRNA. Second, given that inflammation can induce FAHFA synthesis [42], IBMX appears superior over DEX, as DEX has strong anti-inflammatory effects. Finally, we identified DGAT2 [43] and ACSL1, both upregulated selectively by IBMX, as potential contributors to FAHFA synthesis. DGAT2 activity requires acyl-CoA provided by ACSL1, even though it remains to be determined if ACSL1 possess specificity toward FAHFAs. Nevertheless, ACSL1 plays a role in the control of FAO in adipocytes [44], the pathway, which was activated selectively by IBMX. The importance of active FAO for FAHFA synthesis was supported also by the correlation between *PGC1α* and FAHFA content. However, mitochondrial FAO inhibition did not decrease FAHFA levels—and in fact partially mimicked the effect of IBMX—indicating that this positive correlation likely reflects the known link between ATGL activity and FAO [45], rather than a direct causal requirement for oxidation.

The most novel insight into FAHFA synthesis regulation came from using peroxisomal FAO inhibitor TRD. Its ability to enhance IBMX-driven FAHFA levels suggests that synthesis is limited by the availability of precursors (most likely HFAs [46]) normally cleared via peroxisomal FAO. Our data thus reveal a regulatory node at the level of substrate competition: during lipolysis, a significant fraction of HFAs is presumably channeled toward peroxisomal degradation. The fact that blocking this degradative pathway further enhances FAHFA levels indicates that the synthetic machinery is not operating at maximal capacity during lipolysis, limited instead by the availability of free HFAs.

Crucially, TRD alone failed to stimulate synthesis, implying that high HFA availability must be coupled with active lipolysis—specifically ATGL activity—to drive FAHFA flux. Although the partial increase in FAHFAs following ETO treatment suggests that ATGL may not be an absolute requirement if general FFA levels are sufficiently high, it remains the dominant physiological regulator. This “two-hit” requirement (lipolytic supply and HFA availability) reconciles our findings with Santoro et al. [47], who reported stable HFA levels during fasting despite increased FAHFA synthesis. We demonstrate that maximizing production requires the synergy between high lipolytic supply and restricted degradation—a combination not addressed in their study. This dual requirement aligns with physiological states such as exercise (lipolysis coupled with ROS-driven HFA production) and aging, where declining peroxisomal FAO [48]. may explain higher SAHSA levels in older women. Together, our data identify peroxisomal FAO as a new determinant of FAHFA homeostasis.

### 4.6. Limitations

Several limitations of the current study warrant discussion. First, the groups were not matched for FFM, which may have influenced the independent impact of adiposity on the variables studied. Also, our findings are primarily generalizable to sedentary women; sex-based metabolic differences and our focus on untrained individuals may limit extrapolation to men or athletic populations. Second, to minimize participant burden, SAT biopsies were not obtained following the exercise and control sessions. Instead, we prioritized the analysis of circulating FAHFA levels due to their hypothesized direct effects on muscle IS and insulin secretion. Finally, despite utilizing a state-of-the-art targeted lipidomics approach, we were not able to detect 5-PAHSA or other FAHFAs with branching at carbons 2-5, likely reflecting the detection limits of our LC-MS analysis.

### 4.7. Conclusions

In summary, our study provides detailed insights into the regulation of FAHFA levels in women. We demonstrated that age and adiposity differentially impact FAHFA levels as well as the position of FAHFA branching in human SAT, but not in circulation. The lack of correlation between SAT and circulating FAHFA levels challenges the notion of SAT as the primary source of circulating FAHFAs. Furthermore, IS correlated with FAHFA levels in SAT, but not in circulation, suggesting an autocrine/paracrine role for these lipokines within the AT microenvironment.

While circulating FAHFA levels are not affected by acute exercise, our *in vitro* data reveal that FAHFA levels in adipocytes are increased under the conditions leading to high intracellular FFA levels, i.e. when lipolysis is stimulated and/or mitochondrial β-oxidation is inhibited, and further augmented by suppressing the competing peroxisomal FAO pathway.

## Supporting information

STROBE checklist

supplementary table resources

## Data Availability

All data produced in the present study are available upon reasonable request to the authors

## 5. Acknowledgment

We are grateful to Zuzana Varaliová, PhD, for assistance with AT biopsies, and Šárka Fleischerová, for excellent technical help. FAHFA analysis was performed at the Metabolomics Core Facility at the Institute of Physiology of the Czech Academy of Sciences (metabolomics.fgu.cas.cz).

## 6. Author contributions (CRediT)

Conceptualization**: LR, JG, EK, MŠ, MR.** Investigation: **LR, VŠ, JG, MK, MW, MRi, JP, JN, EK, AEH.** Resources: **VŠ, TČ**; Validation: **LR, MRi, TČ, OK, JG**. Formal analysis: **LR, VŠ, MW**. Data curation: **LR, VŠ, MK, MRi, JP.** Writing-original draft: **LR.** Review and editing: **LR, VŠ, JG, MK, MW, MRi, TČ, AEH, JN, OK, MŠ, MR.** Supervision: **LR, TČ, JG**. Visualization **LR, MK, MW**. Project administration: **LR, JG, MR**. Funding acquisition: **LR, JG, TČ, MR**.

## 9. Declaration of generative AI and AI-assisted technologies in the writing process

During the preparation of this work, LR used Google Gemini to edit English language and shorten the text. After using this tool, the authors reviewed and edited the content as needed and take full responsibility for the publication’s contents.

