## supplementary information for "Circulating and Adipose Tissue Profiles of Fatty Acid Esters of Hydroxy-Fatty Acids in Women: Impact of Adiposity, Age, and Acute Exercise"

### Methods and Subjects

This work is based on the clinical study ETAPA (NCT05572905) approved by the Ethical Committee of the Third Faculty of Medicine, Charles University in Prague and the Ethical Committee of Kralovske Vinohrady University Hospital (EK-VP/40/0/2020). The study was conducted between May 2022 and March 2023 at Kralovske Vinohrady University Hospital as a single-center, non-randomized trial. All subjects provided their informed consent before the start of the study. Schemes of study flow and design are shown in Figure 1A and B. In total, 316 Caucasian women (sex assigned at birth) willing to enter the study were evaluated for compliance with inclusion or exclusion criteria. Only one sex, i.e. women, was selected for the study, as we wanted to filter out the possible influence of sex differences that exist at the AT level, especially in response to exercise [1]. The inclusion criteria were age 25–40 and 65–80 years, BMI 18.5–45 kg/m$^2$. The exclusion criteria were diagnosed cancer, type 1 and type 2 diabetes mellitus, liver and renal diseases, major cardiovascular event, bariatric surgery, allergy to lidocaine, positive serology for hepatitis (B and C) and HIV, smoking above 10 cigarettes/day, alcohol consumption above 66g/day, sleep apnea, poor venous status, weight-change more than 3 kg in the last 3 months, untreated hyper- or hypo-thyroidism, and long-term use of medication and/or steroids. Besides that, shift workers and individuals with abnormal sleep/wake patterns were excluded.

A total of 60 women met the criteria and have been assigned to the clinical program. These women were divided into two groups according to their BMI: 1) lean (BMI 18–25, n=20) and 2) obese (BMI 30–40, n=20), and one group according to their age 3) older (age 65-80 years, n=20).  Groups 1 and 2 were matched for age. Last group 4) young (n=20) was selected from groups 1 and 2 to match individually with women from group 3 by % FM assessed by DXA. It included 9 women from group 1 and 11 women from group 2. This approach was used to reduce the bias from aging-associated changes of body composition, when young and older women were matched for % fat mass rather than BMI, which does not change linearly with age. At the end of the trial, one obese woman had to be excluded because of supraphysiological levels of insulin.

Due to the exploratory nature of this study and the lack of established reference ranges for FAHFA levels, a formal power calculation was not performed. The sample size (n=20) is consistent with previous lipidomic studies investigating FAHFA in similar clinical settings [2, 3].

**Baseline clinical investigations and AT sampling**

Anthropometric characteristics were recorded by a trained nurse. Body composition was measured using bio-impedance analysis and Dual emission X-ray absorptiometry. Respiratory quotient was measured with indirect calorimetry and resting metabolic rate was calculated according to modified Weir equation [4]. Glucose tolerance and various IS indexes were assessed by an oral glucose tolerance test (OGTT): after an overnight fast, subjects were asked to drink 75 g of glucose within 5 minutes, blood samples were taken at -15, 0, 30, 60, 90 and 120 minutes.

Needle biopsies of superficial SAT were performed in the abdominal area, 8-10 cm laterally from the umbilicus, after an overnight fast. After disinfection of the area, the skin and AT were anesthetized with 1% mesocaine solution. The skin was incised with a scalpel and a 16G needle with a syringe was used to aspirate AT, then the area was compressed and cooled. The samples of AT were washed with PBS, gently dried, divided into aliquots, snap frozen in liquid nitrogen and then stored in -80°C until analysis.

**Exercise and control intervention**

A week or more prior to any of the two interventions, the maximum oxygen consumption of each participant was assessed during the stress test using bicycle ergometer ($VO_{2max}$). During the stress test, the relative heart rate accompanying the lowest recorded respiratory quotient (RQ) value linked with the maximal oxidation of lipids was determined individually for each woman. This relative heart rate was set up as the target value for the exercise intervention (EXE). The EXE included 60 minutes of aerobic exercise using ergometers at the individually set intensity and a subsequent 60-minute recovery. To ensure the maintenance of the zone of maximal fat oxidation, throughout the cycling, the workload was dynamically adjusted to allow for stable predefined heart rate. To control for the effect of fasting-induced lipolysis, circadian variations, and other variables, the control intervention (CON) involving 2 hours of sitting, with no food and free access to water, was performed in cross-over design. Both EXE and CON interventions were done in the morning after 10 hours of overnight fasting, with no exercise 48 hours prior to the intervention. The order of the interventions was randomly assigned, both interventions were separated from each other, as well as from the day of AT biopsy by at least 1 week. Blood samples were collected immediately before and after the exercise as well as after the recovery period and at the same timepoints during the CON intervention (0, 60, 120 min).

**Plasma/serum analyses and calculations**

Blood insulin and metabolites were analyzed in certified laboratories: glucose was assessed using the hexokinase reaction; insulin by using an electro-chemiluminescent enzyme immunoassay; total cholesterol and triglycerides using enzymatic method kits; high-density lipoprotein-cholesterol (HDL) was measured using polyethylene glycol-modified enzymatic assay kits. Glycerol and FFA were measured using colorimetric assays. Serum protein levels of Serpin Family F Member 1 (Serpin F1), Angiopoietin Like

4 (ANGPTL4), Growth Differentiation Factor 15 (GDF15), and Fatty Acid Binding Protein 4 (FABP4) were analyzed using ELISA DuoSets and Androgen Dependent TFPI Regulating Protein (ADTRP) levels were determined using an ELISA kit.
HOMA-IR, Matsuda index, HIRI and AdipoIR were calculated as summarized by Gastaldelli [5]. HOMA-IR2 and HOMA2-%B were calculated using HOMA2 Calculator v2.2.3 (https://www.dtu.ox.ac.uk/homacalculator/) When possible, average values from three individual days were calculated to lower the impact of day-to-day variability [6].

**FAHFA and FFA analysis**

Samples of AT (100 mg), plasma (100 μL), and *in vitro* cultivated adipocytes (grown in 2 wells of 6 well plate) were extracted according to previously described methods, with several modifications to improve extraction yield and FAHFA detection during LC-MS analysis [7, 8]. Samples were homogenized using a ball mill in a mixture of citric acid buffer and ethyl acetate (ratio 1:2, v/v) containing 1 ng of [$^{13}C_4$]-9-PAHSA as an internal standard. The ethyl acetate phase was collected and dried using a Savant SpeedVac. Based on optimization of extraction yield, we dissolved the dried extracts in 300 μL (200 μL for plasma) of chloroform and loaded the samples onto HyperSep SPE columns (500 mg/10 mL, 40–60 μm, 70 Å) for further purification by solid-phase extraction (SPE). FAHFAs were eluted with 5 mL ethyl acetate and dried. Before analysis, purified extracts were dissolved in 50 μL of a methanol/water mixture (95:5, v/v) to eliminate TGs, potentially suppressing FAHFA signal. Samples were analyzed by LC-MS as before [8]. FFAs in adipocytes were analyzed by untargeted LC/MS using a Vanquish UHPLC System coupled to an Orbitrap Exploris 480 mass spectrometer (Thermo Fisher Scientific, Bremen, Germany). FAHFA extracts were used, as the extraction procedure had been previously validated to fully retain FFA. Data were processed using MS-DIAL software (v4.9.221218) [9], and normalization was performed using locally estimated scatterplot smoothing (LOESS).

**Culture of adipocytes**

Isolation, culture and differentiation of human preadipocytes was described previously [10]. Cells from abdominal SAT from 5 subjects were pooled together, split once and then cryopreserved. For each experiment, a new vial of cells was used to ensure that experiments are performed on standardized cell culture with the same passage number. Cells were plated at the density of 20 000 cells/cm$^2$ and allowed to grow in proliferation medium 4 (DMEM/F12 medium supplemented with 2.5% FBS, 132 nM insulin, 10 ng/mL EGF, 1 ng/mL FGFβ) until two days post-confluence. Differentiation was then induced by DMEM/F12 medium supplemented with 66 nM insulin, 1 μM dexamethasone, 1 nM T3, 0.1 μg/mL transferrin, 0.25 mM IBMX, and 1 μM rosiglitazone. After 6 days, rosiglitazone and IBMX were omitted and dexamethasone was replaced with 0.1 μM cortisol. The differentiation continued until day 12.
On day 12, differentiated adipocytes were twice washed and then treated for 48 hours with either control medium (DMEM/F12 supplemented with 1 nM T3, 10 μg/mL transferrin, and 0.5% FFA and endotoxin-free BSA) or the same medium supplemented with either

1 μM dexamethasone or 250 μM IBMX. Cells were then harvested for the analysis of mRNA and protein levels of selected genes and for FAHFA analysis. In the second experimental set, cells were exposed to 50 μM etomoxir (ETO), 10 μM thioridazine hydrochloride (TRD), or 250 μM IBMX, either alone or in combination (IBMX with ETO or TRD) and harvested for FFA and FAHFA analysis. Both experimental sets included five independent experiments.

**Gene expression analysis**

Total RNA from cells was isolated using PureLink RNA Mini Kit with on column DNAse I treatment. RNA concentration was measured using Nanodrop1000. Total RNA (400 ng) was reverse transcribed using a High-capacity cDNA Reverse Transcription Kit as described previously [11]. For microfluidics, 4 ng of cDNA were preamplified within 18 cycles (SsoAdvanced PreAmpSupermix). For the preamplification, 20x TaqMan gene expression assays of all target genes were pooled together and diluted with water to the final concentration 0.2x for each probe. After preamplification, DNA was diluted 20-times. The qPCR was performed in duplicates on Biomark Real Time qPCR system using 96.96 dynamic array. To analyze acyl-CoA synthetase long chain family member 1 (*ACSL1)* mRNA, qPCR on 4ng of non-amplified cDNA was performed in duplicates on Applied Biosystem 7500Fast instrument. Data were normalized to the mean of the reference gene TATA binding protein (*TBP*). For statistical analysis and presentation $2^{-\Delta Ct}$ or $2^{-\Delta\Delta Ct}$ calculation was used.

**Western blots**

Cells were washed two times with PBS and lysed on ice for 30 minutes in RIPA lysis buffer supplemented with protease and phosphatase inhibitors (Complete, PhoStop). Lysates were then centrifuged for 15 minutes at 15,000x *g*, 4°C. Protein concentrations were determined using the Bicinchoninic Acid Assay. Samples were loaded to a 10% acrylamide minigel and electrotransferred onto the nitrocellulose membrane. Membranes were blocked with 5% BSA. Antibodies against Adipose Triglyceride Lipase, Glyceraldehyde-3-Phosphate Dehydrogenase (GAPDH), Peroxiredoxin 6 (PRDX6), and phosphylated form of Hormone Sensitive Lipase (P-HSL, S563) were from Cell Signaling. Antigen-antibody complexes were detected using corresponding secondary antibodies coupled with horseradish peroxidase and the ECL detection system.

**Statistical analysis**

All datasets were tested for normality or lognormality according to the battery of methods including Shapiro-Wilk, D'Agostino&Pearson, Anderson-Darling and Kolmogorov-Smirnov tests. Non-normally distributed variables (including FAHFA levels in STA and plasma) were log transformed for analysis and back transformed for presentation. Since physiological reference ranges for FAHFA levels are not yet established, no data points were excluded as outliers. All values are described as mean ± standard deviation or standard error of mean as indicated. Baseline characteristics of the groups were analyzed using an unpaired Multiple t-test, with *q* value set to 5%. Effects of the EXE and CON intervention and their comparison between the groups were analyzed by two-way repeated measures ANOVA with Tukey correction to account for multiple comparisons. Effect of interventions on circulating FAHFA were analyzed by paired Multiple t-test, with Q value set to 5%. Analysis of gene expression was performed using principal component analysis (PCA) and variable importance in projection (VIP) using MetaboAnalyst 6.0 [12]. Correlations of nonparametric (gene expression) and log transformed data were expressed by Spearman's, resp. Pearson's correlation coefficient. Significance was set at $p < 0.05$. No imputation methods were used for missing data. All statistical analyses were performed using GraphPad Prism version 10.6 for Windows.
