## Supplementary figures and images for "Circulating and Adipose Tissue Profiles of Fatty Acid Esters of Hydroxy-Fatty Acids in Women: Impact of Adiposity, Age, and Acute Exercise"

### supplementary figure 1

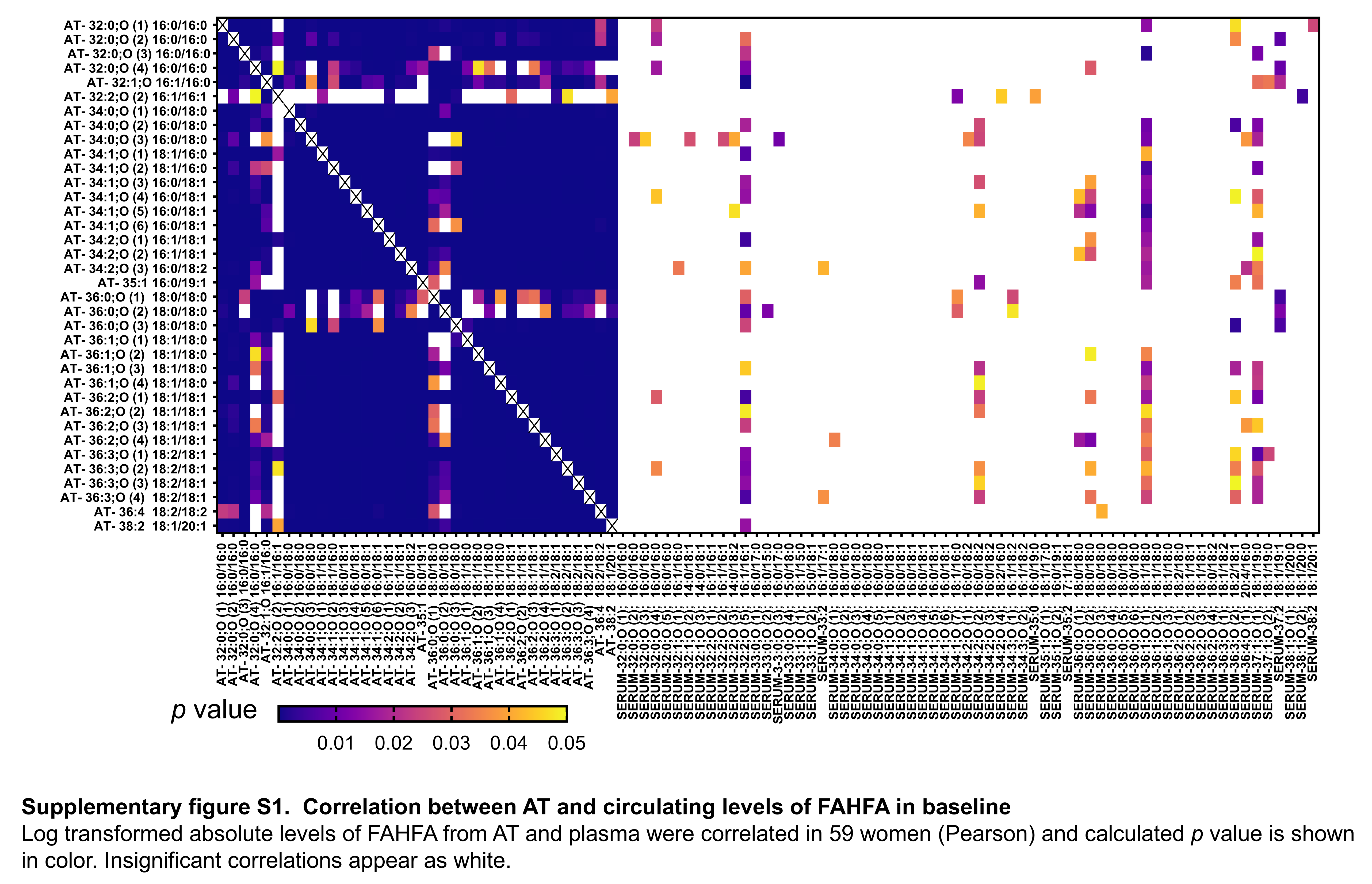
